# Comparison of reported main COVID-19 information sources in national and culturally and linguistically diverse communities in Australia

**DOI:** 10.1101/2021.07.29.21261321

**Authors:** J Ayre, DMM Muscat, O Mac, C Batcup, E Cvejic, K Pickles, H Dolan, C Bonner, D Mouwad, D Zachariah, U Turalic, Y Santalucia, T Chen, G Vasic, K McCaffery

## Abstract

**Background:** To manage the COVID-19 pandemic effectively, governments need clear and effective communication. This is a challenge for culturally diverse communities as groups may have different informational needs and information-seeking behaviours. In this paper we present the frequency of information sources for COVID-19 in a culturally diverse area of Sydney, Australia.

**Methods:** This study reports findings from two surveys. The first recruited participants across 10 languages between March 21 and July 9, 2021. The second provides a point of reference, and was an Australian, nationally-representative sample of English-speaking participants between November 4 – 18, 2020.

**Results:** For the survey in culturally and linguistically diverse communities, of 708 participants, mean age was 45.4 years (SE 0.78), and 51% of respondents were female. Across all language groups, 54.7% of participants used Australian official or public broadcasters to find out about COVID-19 (n=421). Australian commercial information sources (54.1%, n=417), social media (51.6%, n=397), and family and friends in Australia (32.7%, n=252) were common sources. Patterns varied substantially across language groups. In the nationally representative survey (n=2313), 67% of participants (n=1540) used Australian official or public broadcasters, with lower proportions for social media (31.9%, n=738) and friends, family or other personal sources (23.1%; n=533).

**Conclusion:** Almost 50% of participants from culturally and linguistically diverse communities did not report using Australian official or public broadcaster as main sources of information. Instead Australian commercial information sources, friends and family, overseas sources and social media were common. Though a crude comparison of the two datasets, this data can guide policy decisions for communication to different community groups. Further analysis is needed to interpret this data. Better understanding of how diverse communities seek and receive COVID-19 health information is imperative as we manage the current COVID-19 outbreak in the Sydney region.

---

High uptake of COVID-19 prevention behaviours such as distancing, mask-wearing, testing and vaccination are key aspects of pandemic management in Australia. Whilst these behaviours can be mandated through government policy, and opposing behaviours discouraged through fines and other penalties, ultimately the uptake of these behaviours relies on clear and effective public health communication to ensure a high level of understanding, acceptance, and engagement within the community(1).

However, over the course of the COVID-19 pandemic, studies have consistently found that most public health COVID-19 information is written at a level much higher than is recommended for general audiences, let alone for people with low literacy or health literacy (2, 3). For example, a recent study showed that for resources collected in April 2021, the median grade reading score for Australian government COVID-19 information on vaccination, mask-wearing, and distancing ranged from Grade 12 to a postgraduate level (Grade 14)(2), 4-6 grades beyond the recommended Grade 8 reading level (4). Perhaps unsurprisingly, our analysis of over 4000 Australians at the start of the pandemic (April 2020) found that that people with inadequate health literacy and people who did not speak English as their main language at home experienced greater challenges in understanding government COVID-19 information compared to people with adequate health literacy or English as their primary language(5).

Public health efforts must also use communication channels that the community can and do access(6). Our research identified that in April 2020, most participants obtained information about COVID-19 from public television (68%), social media (64%), and government websites (64%)(5). Participants with inadequate health literacy in this survey also reported that information about COVID-19 was more difficult to find. Identifying the most effective communication channels is particularly important for disadvantaged groups, including people from culturally and linguistically diverse communities (6). These channels provide an important mechanism through which tailored messages that are co-created with the community can effectively reach their target audience(6).

Identifying the most appropriate communication channels becomes more challenging in areas with high cultural diversity, as each group may have distinct informational needs and information-seeking behaviours. Areas such as Western and Southwestern Sydney, New South Wales (NSW), are home to dozens of cultural and language groups, with up to 39% of residents born overseas in non-English speaking countries (7). It is therefore critical that we identify strategies to enhance COVID-19 communication by improving the accessibility of existing channels for these groups, or leveraging communication channels that these groups already use. In this paper we present the frequency COVID-19 information sources in 10 language groups across three adjoining regions in Sydney with high cultural diversity: Western Sydney, Southwestern Sydney, and Nepean Blue Mountains, between March and July 2021. Findings were contrasted against those of an Australian nationally representative sample (data collected November 2020).

## Methods

### Culturally and linguistically diverse community survey

#### Study Design

An online survey was conducted with a cross-sectional sample using the web-based survey platform Qualtrics. The study was approved by Western Sydney Local Health District Human Research Ethics Committee (Project number 2020/ETH03085).

#### Setting

Participants were recruited from 21^st^ March to July 9th, 2021. During this period, the COVID-19 vaccines had begun rollout across Australia, and daily cases in NSW ranged from 0 – 46.

#### Participants

Through iterative discussions with multicultural health staff, we selected ten language groups that would provide broad coverage across language groups from different global regions, with varying average levels of English language proficiency (based on 2016 Australian census data), varying access to translated materials, and varying degrees of reading skill in their main language spoken at home. Participants were eligible to take part if they were aged 18 or over and spoke one of the following as their main language at home: Arabic, Assyrian, Croatian, Dari, Dinka, Hindi, Khmer, Mandarin, Samoan, Tongan, or Spanish. Each of the language groups selected was an important group within the Greater Western Sydney region (Western Sydney Local Health District, South Western Sydney Local Health District, Nepean Blue Mountains Local Health District).

Participants were recruited through multicultural and bilingual staff and health care interpreter services staff. Multicultural and bilingual staff recruited participants through their existing networks, community events and community champions. Health care interpreter staff recruited patients at the end of a medical appointment that included an interpreter staff. Potential participants were offered two means of taking part: completing the survey themselves online (English or translated), or completing the survey with assistance from bilingual staff or an interpreter (by phone). To ensure consistency in the phrases used for assisted survey completion, translated versions of the survey were provided to the bilingual staff and interpreters.

#### Survey design

Surveys were available in English or translated. Demographic survey items included age, gender, education, whether born in Australia, years living in Australia, main language spoken at home, English language proficiency, reading proficiency in language spoken at home, access to the internet, smartphones, chronic disease, and a single-item health literacy screener(8). Participants were asked to specify through multiple choice questions their top 3 information sources for finding out about COVID-19 in the previous 4 weeks, via 8 categories (TV, radio, social media, websites, printed materials, ‘family, friends and community’, health professionals, and other. Participants were then asked for more specific answers (e.g. which kind of social media). Survey items also asked about which country overseas information came from and the language it was provided in. Participants were asked to report on perceived difficulty finding easy-to-understand information about COVID-19, both in English and in their main language. Other survey items not reported here included COVID-19 knowledge, attitudes, behaviours, and items capturing the impact of the pandemic.

#### Analysis plan

Within each language group, frequencies of responses were weighted to reflect population (census data) gender and age group distributions (18-29 years, 30-49 years, 50-69 years, ≥70 years). A single participant indicated their gender as ‘other’ and was unable to be included in weighted analyses.

Total recruitment for the Spanish language group were low (<50), with notable gaps for some age groups. For this reason, results for this language group are not presented in the analysis, but are included in total counts. Responses to survey items about information sources in the survey for culturally and linguistically diverse groups were re-coded to reflect the categories presented in Table 1, to facilitate a more meaningful interpretation of the results.

**Table 1.** Main information sources for finding out about COVID-19, by language group, 20 Mar - 9 Jul 2021.

| Language group | Total | Official /public Australian source<sup>a</sup> |  | Australian Commercial<sup>b</sup> |  | Community (local)<sup>c</sup> |  | Friends/ family in Australia<sup>d</sup> |  | Overseas sources<sup>e</sup> |  | Social media |  | Facebook |  | Instagram |  | Whatsapp |  | Youtube |  | other social media |  |
| --- | --- | --- | --- | --- | --- | --- | --- | --- | --- | --- | --- | --- | --- | --- | --- | --- | --- | --- | --- | --- | --- | --- | --- |
|  |  | N | % | N | % | N | % | N | % | N | % | N | % | N | % | N | % | N | % | N | % | N | % |
| Arabic | 80 | 39 | 48.6 | 39 | 48.6 | 5 | 6.8 | 15 | 18.2 | 10 | 12.6 | 54 | 68.1 | 44 | 55.0 | 11 | 13.7 | 22 | 27.1 | 24 | 29.9 | 3 | 3.6 |
| Assyrian | 133 | 77 | 57.8 | 84 | 63.3 | 30 | 22.8 | 43 | 32.4 | 11 | 8.6 | 55 | 41.0 | 50 | 37.3 | 16 | 12.1 | 2 | 1.8 | 21 | 16.1 | 8 | 5.7 |
| Croatian | 121 | 73 | 60.4 | 56 | 46.6 | 79 | 65.3 | 77 | 63.3 | 119 | 98.4 | 46 | 38.3 | 45 | 37.3 | 23 | 18.8 | 40 | 32.9 | 42 | 34.8 | 16 | 13.5 |
| Dari | 44 | 30 | 67.2 | 17 | 38.7 | 3 | 7.7 | 24 | 53.7 | 12 | 28.0 | 32 | 73.2 | 31 | 69.9 | 12 | 26.7 | 4 | 8.5 | 21 | 48.7 | 16 | 37.0 |
| Dinka | 63 | 42 | 66.5 | 35 | 54.8 | 16 | 25.1 | 9 | 14.6 | 3 | 4.9 | 41 | 65.2 | 35 | 55.3 | 15 | 23.7 | 12 | 19.5 | 16 | 25.2 | 18 | 28.8 |
| Hindi | 42 | 29 | 68.5 | 31 | 73.7 | 9 | 21.9 | 6 | 15.2 | 14 | 32.8 | 30 | 71.1 | 19 | 44.5 | 13 | 30.0 | 19 | 46.0 | 15 | 35.5 | 6 | 13.9 |
| Khmer | 63 | 58 | 91.7 | 52 | 81.8 | 22 | 35.4 | 21 | 32.6 | 3 | 5.3 | 36 | 56.6 | 34 | 54.5 | 26 | 42.0 | 5 | 7.3 | 18 | 29.3 | 17 | 26.6 |
| Chinese | 76 | 34 | 44.2 | 35 | 46.1 | 14 | 18.7 | 35 | 46.7 | 25 | 33.1 | 54 | 71.5 | 11 | 14.6 |  | 0.0 | 5 | 6.3 | 19 | 24.7 | 40 | 52.5 |
| Samoan/Tongan | 42 | 12 | 28.9 | 34 | 81.9 | 12 | 27.5 | 7 | 16.7 | 3 | 6.6 | 21 | 49.0 | 17 | 40.3 | 8 | 19.0 | 1 | 1.5 | 5 | 11.5 | 42 | 100.0 |
| **Total** | **770** | 421 | 54.7 | 417 | 54.1 | 196 | 25.5 | 252 | 32.7 | 205 | 26.7 | 397 | 51.6 | 312 | 40.5 | 136 | 17.7 | 112 | 14.5 | 189 | 24.5 | 124 | 16.1 |

<sup>a</sup> Official Australian / Australian public broadcaster sources: Government websites, health professionals, Public TV, public radio/podcast
<sup>b</sup> Commercial Australian sources: commercial TV, commercial radio/podcast, newspapers
<sup>c</sup> Community: community TV, community radio/podcast, community leader, religious leader
<sup>d</sup> Family/friends living in Australia
<sup>e</sup> Overseas: TV, radio/podcast, government/news website, family/friends who live overseas

**Table 2.**
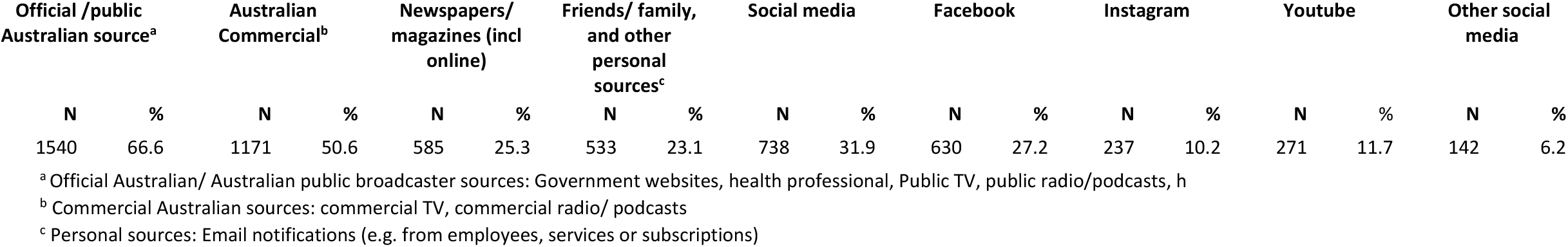
Main information sources for finding out about COVID-19 from nationally representative Australian survey, Nov 2020.

| Official /public Australian source<sup>a</sup> |  | Australian Commercial<sup>b</sup> |  | Newspapers/ magazines (incl online) |  | Friends/ family, and other personal sources<sup>c</sup> |  | Social media |  | Facebook |  | Instagram |  | Youtube |  | Other social media |  |
| --- | --- | --- | --- | --- | --- | --- | --- | --- | --- | --- | --- | --- | --- | --- | --- | --- | --- |
| N | % | N | % | N | % | N | % | N | % | N | % | N | % | N | % | N | % |
| 1540 | 66.6 | 1171 | 50.6 | 585 | 25.3 | 533 | 23.1 | 738 | 31.9 | 630 | 27.2 | 237 | 10.2 | 271 | 11.7 | 142 | 6.2 |

<sup>a</sup> Official Australian/ Australian public broadcaster sources: Government websites, health professional, Public TV, public radio/podcasts, h
<sup>b</sup> Commercial Australian sources: commercial TV, commercial radio/ podcasts
<sup>c</sup> Personal sources: Email notifications (e.g. from employees, services or subscriptions)

### Nationally representative survey of Australian adults (November 2020)

#### Study Design

An online survey was conducted with a cross-sectional sample using the web-based survey platform Qualtrics. This study was approved by the University of Sydney Human Research Ethics Committee (2020/212).

#### Setting

The survey was distributed Australia-wide between 4-18 November 2020, when restrictions were considerably eased across Australia as no locally acquired cases were recorded for the first time since June 2020.

#### Participants

Participants were aged 18 years and older, able to read and understand English, and currently residing in Australia. Participants were recruited via Dynata, who have more than 600 000 online Australian panel members aged older than 18 years who have consented to participate in online research. Panel members were sent an email invitation to participate in the study and received points for completing the survey, which they could redeem for gift vouchers, donations to charities, or money. We purposively set quotas to recruit a nationally representative sample by age, gender and education.

#### Survey design

The survey was only available in English. Demographic items were similar to those reported in a previous Australia-wide survey, reported elsewhere (5), and survey items covered a broad range of topics, including knowledge about COVID-19, information sources, attitudes, behaviours and the pandemic’s impact on individuals. Participants were asked to specify through multiple choice questions their top 3 information sources for finding out about COVID-19.

#### Analysis plan

Descriptive statistics were used to summarise demographic characteristics and information sources in SPSS Version 25.

## Results

### Culturally and linguistically diverse community survey

A total 708 respondents were recruited (442 (62.4%) self-completed, 266 (37.6%) received assistance through an interpreter). The mean age was 45.4 years (standard error [SE] 0.78; range 18–91 years), and 51% of respondents were female. Most participants (88%) were born in a country other than Australia; 31% reported that they did not speak English well or at all; 70% had no tertiary qualifications. Inadequate health literacy was identified for 41% of the sample.

Across all language groups, more than half of participants used Australian official or public broadcaster information sources (including TV, radio, websites, and health professionals) (54.7%, n=421), Australian commercial information sources (54.1%, n=417), and social media (51.6%, n=397), as their main ways of finding out about COVID-19 (Table 1). Around one third of participants stated that friends or family in Australia were a main information source (32.7%, n=252). Around one quarter of participants responded that a main way of finding out about COVID-19 was through community (including community and religious leaders, community TV and radio) (25.5%, n=196), or from information sources from overseas (including TV, radio, websites, and family or friends living overseas) (26.7%, n=205). Almost half (44.5%, n=312) indicated that these sources were mostly in another language other than English. For participants who indicated that they used overseas sources, 81% also responded that their information was mostly communicated in a language other than English.

Patterns varied within language groups (Table 1). For example, official and public broadcasters were used least by Samoan/Tongan speakers (28.9%), and most by Khmer speakers (91.7%). Community information sources were most commonly identified for Croatian speakers (65.3%), followed by Khmer speakers (35.4%); reliance of friends and family living in Australia was highest for Croatian speakers (63.6%), followed by Dari speakers (53.7%), and Chinese speakers (46.7%). The language groups that ranked overseas information sources highly included Croatian (98.4%), Chinese (33.1%), Hindi (32.8%), and Dari (28.0%). Chinese speakers reported the highest rate of using social media to find out about COVID-19 (71.5%). More than half of Chinese speakers reported using WeChat (51.5%) to find out about COVID-19; 24.5% reported using Youtube).

### Nationally representative survey

A total of 2313 respondents were recruited for the nationally representative survey. The mean age was 45.1 years (SD=16.9 years) and 47.6% were female. 29.7% (n=688) resided in New South Wales,23.6% in Victoria, and 19.7% in Queensland. The majority (77.6%) had adequate health literacy as assessed by the Single Item Literacy Screener and 61.4% had tertiary qualifications. 5.8% reported speaking a language other than English at home and 4.2% identified as Aboriginal or Torres Strait Islander.

Two thirds of participants recruited for the nationally representative survey used Australian official or public broadcaster information sources for COVID-19 (including TV, radio, websites, and health professionals) (66.6%, n=1540) and Australian commercial information sources (50.6%, n=1171) as their main ways of finding out about COVID-19. Around one third of participants stated that social media was a main information source (31.9%, n=738); 25.3% (n=585) reported using newspapers and/or magazines and 23.1% used personal and other sources (n=533).

## Discussion

This study found that for a survey of COVID-19 information across 10 language groups in Sydney conducted between March and July 2021, almost 50% of participants did not report using Australian official or public broadcaster as main sources of information. Instead Australian commercial information sources, friends and family, overseas sources and social media were common. In this paper we present simple counts and percents for each cultural group. A more comprehensive statistical analysis is currently underway. We present a crude comparison with data collected from our nationally representative sample of Australian adults collected in November 2020 to illustrate differences and similarities between information sources to guide policy decisions for communication to different community groups. In the Australian national sample 67% of adults reported official sources and the public broadcaster as their main official source. Further analysis is needed to interpret this data fully and is currently underway.

We highlight, the observed pattern of information sources differed substantially across language groups. This reiterates the importance of tailored public health communication, including the communication channels used. Better understanding of how diverse communities seek and receive COVID-19 health information is imperative as we move forward with successful roll out of COVID-19 vaccinations and as we manage the current COVID-19 outbreak in the Sydney region.

## Data Availability

Data available upon reasonable request from corresponding author.

